# The course of COVID-19 in allergic rhinitis patients receiving allergen spesific immunotherapy

**DOI:** 10.1101/2022.01.29.22270072

**Authors:** Emel Atayik, Gökhan Aytekіn

**Author notes:** **Correspending authors Address for reprints: Emel Atayik**, Allergy and Clinical İmmunology Specialist, University of Health Sciences Konya City Hospital, Division of Allergy and Clinical Immunology, Konya / TURKEY. **Informed consent** The study protocol was approved by the Ethics committee of the Karatay University (with the Meeting number: 9, decision number: 2021/036). Manuscripts that have not been presented orally or as a poster. **Financial disclosure:** The authors have no financial relationship relevant to this article to disclose. **Disclaimers:** None.

## Abstract

**Introduction:** The aims of presenting study were trying to expose the course of SARS-CoV-2 (severe acute respiratory syndrome-related coronavirus) in patients with allergic rhinitis (AR), to compare the prevalence of SARS-CoV-2 infection, hospitalization and pneumonia rates in patients with AR receiving allergen immunotherapy (AIT) and patients did not receive AIT (non-receivers) and to define possible risk factors for SARS-CoV-2 positivity in patients with AR.

**Materials and Methods:** A total of 419 patients with AR who were being followed up in a tertiary allergy clinic between 1 June 2020 and 31 December 2020, were selected for the study. Only patients who were receiving active-continuous treatment for allergic rhinitis during the study period, were included in the study.

**Results:** Seventy-nine patients (18.9%) became infected with the SARS-CoV-2 [32 patients (19.6%) in AR patients with AIT and 47 patients (19.0%) in non-receivers] and the rate of pneumonia was 2.4% [12.7% of SARS-CoV-2 (+) patients]. There was no significant difference was determined between the AR patients with AIT and the non-receivers in regard to the rate of SARS-CoV-2 infection, pneumonia, and hospitalization (p: 0.864, p: 0.081, p: 0.113). There was a significant difference between the groups in terms of gender, duration of disease, sensitivity to allergens (atopy), and serum IgE levels (p: 0.009, p: 0.001, p: 0.001, and p: 0.001). The accompanying comorbidities, eosinophil count, AIT, and duration of AIT were not found to be associated with an increased risk SARS-CoV-2 PCR positivity. However, the female gender was shown to be associated with a decreased risk for SARS-CoV-2 PCR positivity (OR, 0.571; 95% confidence interval, 0.330-0.987; p: 0.045)

**Conclusion:** The course of SARS-CoV-2 is similar in patients with AR who underwent AIT and patients with AR who did not undergo AIT, and AIT does not seem to increase the risk for SARS-CoV-2 infection.

## Introduction

Although unprecedented efforts have been made around the world to prevent the spread of COVID-19 (Coronavirus disease 2019) and contain the disease, the number of COVID-19 cases continues to increase, and since it was first identified, COVID-19 has become the most important health problem all over the world (1, 2). There is still no effective treatment for the disease, and due to different virus variants and various socioeconomic inequalities, vaccination studies against the virus do not have the expected speed and effect for now. Therefore, both in terms of reducing mortality and morbidity, it is very important to identify special patient groups, especially those with chronic diseases and who may be more affected by COVID-19, and to investigate the effect of the treatment received by these patient groups on the course of COVID-19, and efficient utilization of resources.

Allergic rhinitis (AR) is a common allergic disease that affects approximately 10-30% of the pediatric and adult population (3, 4). Allergen Immunotherapy (AIT) can be applied to patients with AR who do not have sufficient clinical benefit despite minimal allergen exposure and optimal pharmacological treatment or who have side effects related to these treatments. Among patients with AR, AIT can be applied to those who do not have sufficient clinical benefit despite minimal allergen exposure and optimal pharmacological treatment or who have side effects related to these treatments. Patients with AR tend to produce lower levels of type 1 interferon (IFN) during upper respiratory viral infections than those without AR (5, 6). This may put allergic patients at higher risk for COVID-19-related morbidity and mortality (7, 8). In contrast, low expression of angiotensin converting enzyme (ACE)2 was detected in airway cells of AR patients, and this is thought to be protective against COVID-19.

In patients with seasonal/perennial allergic rhinitis, AIT is the only therapy with disease-modifying and therapeutic potential and induces long-term tolerance of the immune system to allergens. AIT exerts these effects with an increase in B regulatory (Breg) and T regulatory (Treg) cell numbers and functions, and IL-10 levels in the foreground. (9, 10). AIT inhibits the activation of allergen-specific Th2 cells through B and T reg cells, as well as suppresses the T cell response directed by Th1 and Th17 cells. Besides causing cytokine storm of dysregulated Th1 immune response and ARDS (Acute respiratory distress syndrome), ARDS is an important cause of SARS-CoV-2 related mortality (11). Therefore, it can be argued that AIT may have a positive effect on the course of SARS-CoV-2. Moreover, adenomatous and secretory structure of the nasal mucosa in AR patients due to allergen exposure, local anti-inflammatory effects of intranasal steroids used for treatment, type 2 inflammation dominance in AR and the effects of AIT on Breg and Treg suggest that the course of SARS-CoV-2 may differ in patients with AR.

Therefore, the aims of presenting study were trying to expose the course of SARS-CoV-2 in patients with allergic rhinitis, to compare the prevalence of SARS-CoV-2/COVID-19 infection, hospitalization and mortality rates in patients with allergic rhinitis receiving AIT and patients did not receiving AIT (non-receivers) and to define possible risk factors for SARS-CoV-2 positivity in patients with allergic rhinitis.

## Material and Methods

Four hundred nineteen adult patients with allergic rhinitis, who were being followed-up in a tertiary allergy clinic in Konya, located in the central Anatolia, Turkey, between June 2020 and December 31, 2020, were selected for the study. Only patients who were receiving active-continuous treatment for allergic rhinitis during the study period, were included in the study.

Demographic (age, gender, duration of allergic rhinitis and AIT, accompanying comorbidities, atopy) and clinical data (serum IgE and blood eosinophil counts in patients prior to initiation of treatment for allergic rhinitis and/or AIT) were retrieved from medical files. Skin prick test was performed by with standardized inhalant allergens (ALK, Madrid, Spain) House dust mite (Dermatophagoides (D) farinae, D. pteronyssinus), cat (Felis domesticus), dog (Canis familaris), cockroach (Blatella germanica), fungi (Alternaria, Cladosporium, Aspergillus), and pollen mixtures (tree, weed, grass) were used for prick test. Specific IgE measurement was carried out in all patients in which systemic atopy could not be demonstrated with skin prick tests. It was performed subcutaneously with the conventional protocol in all patients undergoing AIT. We did not have any patients who underwent sublingual immunotherapy, rush or ultra-rush immunotherapy. The diagnosis of SARS-CoV-2 was made by a positive Polymerase Chain Reaction (PCR) test in patients with consistent clinical presentation for COVID-19.

Abbott Cell Dyn 3700 series (Sheath reagent) and Siemens BN II/ BN ProSpec system (using particle-enhanced immunonephelometry) were used for whole blood count and quantitative determination of serum immunoglobulin (Ig) E.

The study was approved by Karatay University Ethics Committee (Decision number 2021/36, date 19.11.21)

Statistical analysis was performed with the IBM SPSS Statistics Version 22 software package. Normally distributed parameters were presented as mean ± standard deviation and data that were not normally distributed were expressed as median (interquartile range: minimum– maximum). Descriptive data were presented as frequencies and percentages and compared using a Chi-square test. Comparisons between baseline characteristics were performed by independent Student t, Mann-Whitney rank-sum, Fisher exact or Chi-square tests where appropriate. As a result of these statistical analysis, parameters with p<0.2 between SARS-CoV-2 (+) patients and SARS-CoV-2 (-) patients were subjected to regression analysis. Binomial logistic regression analysis was performed to determine independent predictors for SARS-CoV-2 positivity.

## Results

A total of 419 patients with allergic rhinitis were included in the study [Female: 256 (63.5%), Male: 163 (36.5%)]. The mean age was 30 years (18 to 76 years). The mean duration of disease was 5 years (0.6-35). In patients undergoing AIT, the duration of immunotherapy was 15 months (3-58 months). The most common allergen sensitivity in patients was found to be pollen mixture sensitivity (58.7%) and house dust mite sensitivity (23.2%). Mean serum IgE level was 90 (10-2020) IU/ml and eosinophil count was 170 (3.40-1100) cells/ml. One hundred sixty-three patient (38.9%) were receiving AIT.

Seventy-nine patients (18.9%) became infected with the SARS-CoV-2 virus during the study period [32 patients (19.6%) in allergic rhinitis patients with AIT and 47 patients (19.0%) in non-receivers]. There was no significant difference was determined between the allergic rhinitis patients with AIT and the non-receivers in regard to the rate of SARS-CoV-2 (p: 0.864). Clinical characteristics of the patients are summarized in Table 1.

**Table 1:** Demographic, clinical and laboratory parameters of allergic rhitis patients

| Parameters | Total<br>(n: 419) | Allergic rhinitis<br>patients with AIT<br>(n: 163) | Allergic rhinitis<br>patients with out AIT<br>(n: 256 ) | p |
| --- | --- | --- | --- | --- |
| Age, years | 30 (18-76) | 31 (18-69) | 28 (18-76) | 0.070 |
| Gender, Female, n % | 256 (63.5) | 91 (55.8) | 175 (68.4) | **0.009** |
| Duration of disease, years | 5 (0.6-35) | 6 (1-35) | 4 (0.6-24) | **0.001** |
| Allergen sensivity, n (%) |  |  |  | **0.001** |
| • Pollen mixtures | 246 (58.7) | 115 (70.6) | 131 (51.2) | **0.001** |
| • House dust mite | 97 (23.2) | 41 (25.2) | 56 (21.9) | 0.438 |
| • Mold | 6 (1.4) | 0 | 6 (2.3) | 0.086 |
| • Animal dander | 7 (1.7) | 1 (0.6) | 6 (2.3) | 0.178 |
| • Venom | 20 (4.8) | 3 (1.8) | 17 (6.6) | **0.032** |
| • Multiple | 43 (10.3) | 3 (1.8) | 40 (15.6) | **0.001** |
| IgE, at diagnosis, IU/ml | 90 (10-2020) | 118 (17-2020) | 78 (10-1143) | **0.001** |
| Eosinophil count, cell/ml | 170 (3.40-1110) | 160 (3.40-1110) | 170 (10-980) | 0.971 |
| SARS-CoV-2, n % | 79 (18.9) | 32 (19.6) | 47 (19) | 0.864 |
| Pneumonia, n % | 10 (2.4) | 7 (4.3) | 3 (11.5) | 0.300 |
| Hospitalization | 7 (1.7) | 5 (15.6) | 2 (7.7) | 0.356 |

AIT: Allergen immunotherapy, Ig: immunoglobulin, SARS-CoV-2: Severe acute respiratory syndrome coronavirus 2

Within the 419 patients included in the study, the rate of pneumonia was 2.4% (10 patients) [12.7% of SARS-CoV-2 (+) patients]. During the study period, seven patients [21.9% of SARS-CoV-2 (+) patients] in AIT group and three patients [6.4% of SARS-CoV-2 (+) patients] in non-AIT group had pneumonia due to SARS-CoV-2. Five patients [15.6% of SARS-CoV-2 (+) patients] in AIT group and two patients [4.3% of SARS-CoV-2 (+) patients] in non-AIT group were hospitalized. There was no significant difference was determined between allergic rhinitis patients with AIT and the non-receivers in regard to the rate of pneumonia and hospitalization (p: 0.081 and p: 0.113). There were no patients admitted to the intensive care unit or died during the study period.

We divided the study participants into two group as the allergic rhinitis patients on AIT and allergic rhinitis patients not receiving AIT; no significant difference was determined between the groups in terms of age, baseline eosinophil count and frequency of infection with SARS-CoV-2 virus, SARS-CoV-2 related pneumonia and SARS-CoV-2 related hospitalization.

However, there was a significant difference between the groups in terms of gender, duration of disease, sensitivity to allergens (atopy) and serum IgE levels (p: 0.009, p: 0.001, p: 0.001 and p: 0.001, respectively). (Table 1)

When SARS-CoV-2 positive and negative allergic rhinitis patients were compared; there were no significant differences between both groups in terms of age, duration of disease, accompanying comorbidities, sensitivity to allergens (atopy), serum IgE levels, eosinophil counts, rate of patients receiving AIT and duration of AIT. A significant difference was determined in terms of gender (p: 0.042) (Table 2).

**Table 2:**
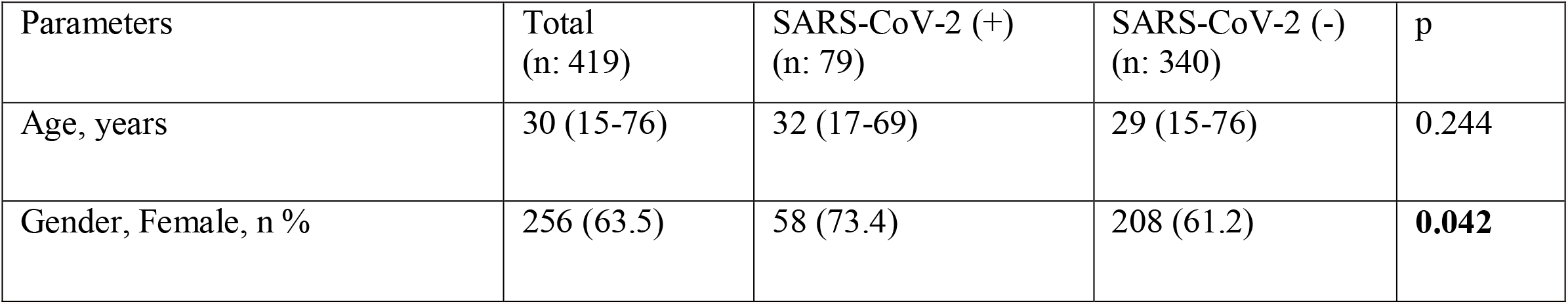
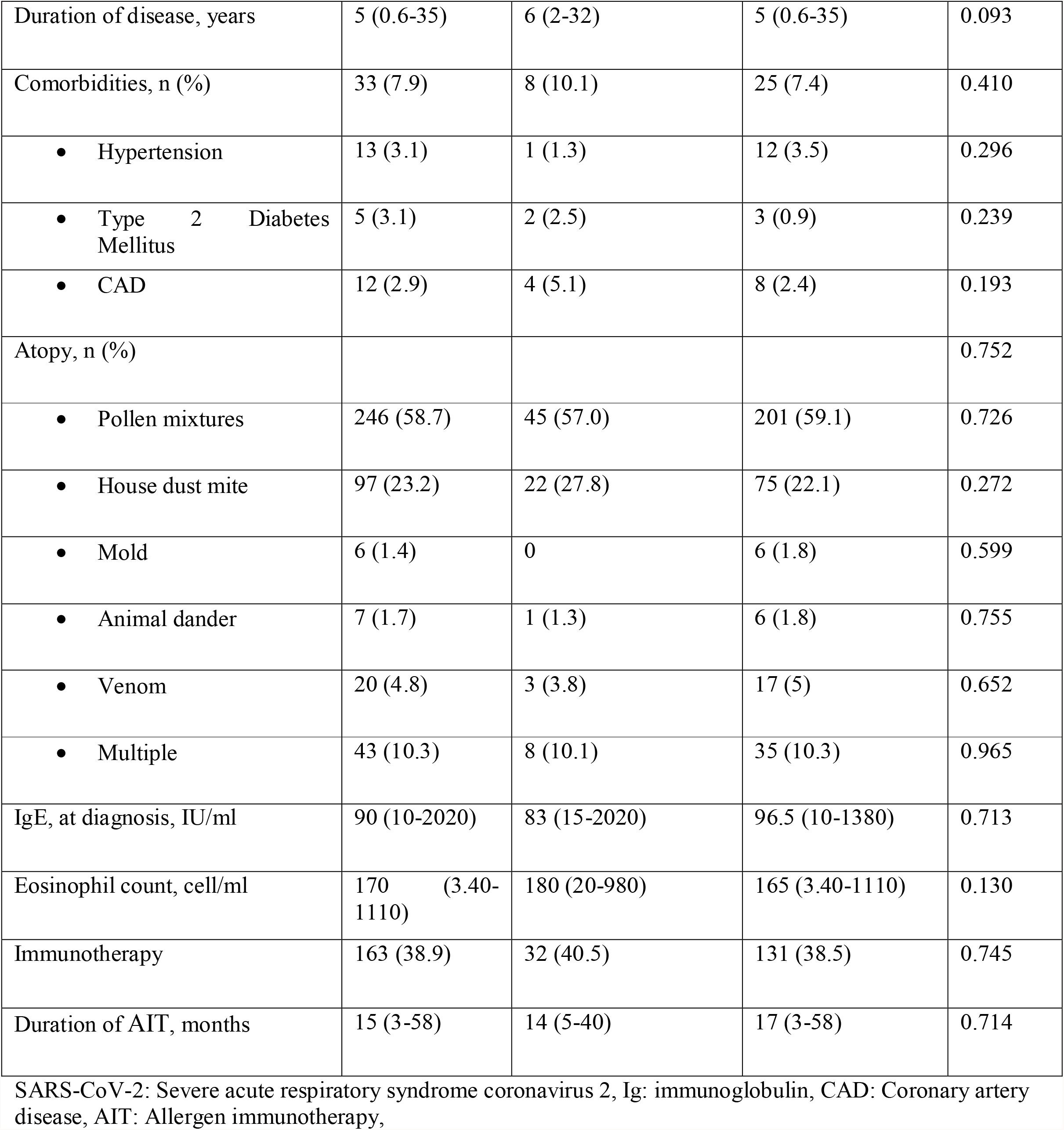
Demographic and clinical characteristics of allergic rhinitis patients according to SARS-CoV-2

| Parameters | Total<br>(n: 419) | SARS-CoV-2 (+)<br>(n: 79) | SARS-CoV-2 (-)<br>(n: 340) | p |
| --- | --- | --- | --- | --- |
| Age, years | 30 (15-76) | 32 (17-69) | 29 (15-76) | 0.244 |
| Gender, Female, n % | 256 (63.5) | 58 (73.4) | 208 (61.2) | **0.042** |
| Duration of disease, years | 5 (0.6-35) | 6 (2-32) | 5 (0.6-35) | 0.093 |
| Comorbidities, n (%) | 33 (7.9) | 8 (10.1) | 25 (7.4) | 0.410 |
| • Hypertension | 13 (3.1) | 1 (1.3) | 12 (3.5) | 0.296 |
| • Type 2 Diabetes Mellitus | 5 (3.1) | 2 (2.5) | 3 (0.9) | 0.239 |
| • CAD | 12 (2.9) | 4 (5.1) | 8 (2.4) | 0.193 |
| Atopy, n (%) |  |  |  | 0.752 |
| • Pollen mixtures | 246 (58.7) | 45 (57.0) | 201 (59.1) | 0.726 |
| • House dust mite | 97 (23.2) | 22 (27.8) | 75 (22.1) | 0.272 |
| • Mold | 6 (1.4) | 0 | 6 (1.8) | 0.599 |
| • Animal dander | 7 (1.7) | 1 (1.3) | 6 (1.8) | 0.755 |
| • Venom | 20 (4.8) | 3 (3.8) | 17 (5) | 0.652 |
| • Multiple | 43 (10.3) | 8 (10.1) | 35 (10.3) | 0.965 |
| IgE, at diagnosis, IU/ml | 90 (10-2020) | 83 (15-2020) | 96.5 (10-1380) | 0.713 |
| Eosinophil count, cell/ml | 170 (3.40-1110) | 180 (20-980) | 165 (3.40-1110) | 0.130 |
| Immunotherapy | 163 (38.9) | 32 (40.5) | 131 (38.5) | 0.745 |
| Duration of AIT, months | 15 (3-58) | 14 (5-40) | 17 (3-58) | 0.714 |

SARS-CoV-2: Severe acute respiratory syndrome coronavirus 2, Ig: immunoglobulin, CAD: Coronary artery disease, AIT: Allergen immunotherapy,

It was found that, according to univariant and multivariant regression analysis, the accompanying comorbidities (coronary artey disease), eosinophil count, AIT and duration of AIT were not found to be associated with an increased risk SARS-CoV-2 PCR positivity. On the other hand, female gender was shown to be associated with an decreased risk for SARS-CoV-2 PCR positivity (OR, 0.571; 95% confidence interval, 0.330-0.987; p: 0.045) (Table 3)

**Table 3.** Logistic regression analysis of possible risk factors associated with SARS-CoV-2 in allergic rhinitis patients

| Variables | Univariate Analysis |  | Multivariate Analysis |  |
| --- | --- | --- | --- | --- |
|  | OR (95% CI) | P value | OR (95% CI) | P-value |
| **Gender** | 0.571 (0.331-0.984) | **0.043** | 0.571 (0.330-0.987) | **0.045** |
| **Duration of disease** | 1.041 (1.000-1.083) | 0.048 | 1.040 (0.999-1.082) | 0.056 |
| **CAD** | 2.213 (0.649-7.543) | 0.204 |  |  |
| **Eosinophil count** | 1.001 (1.000-1.002) | 0.073 |  |  |
| **Immunotherapy** | 1.086 (0.659-1.790) | 0.745 |  |  |
| **Duration of immunotherapy** | 0.987 (0.953-1.022) | 0.470 |  |  |

## Discussion

To the best our knowledge, our study is the only study evaluating the course of COVID-19 in patients undergoing AIT, and this study highlights 3 important findings (1). Allergic disease duration, sensitized allergen types, and IgE levels at the time of diagnosis are higher in allergic rhinitis patients undergoing AIT (2). There is no significant difference between the patients who underwent AIT and the non-AIT patient group in terms of SARS-CoV-2 prevalence, SARS-CoV-2-associated pneumonia, and SARS-CoV-2-associated hospitalization (3). Although female gender is protective for SARS-CoV-2 positivity in allergic rhinitis patients, AIT or duration of AIT is not a risk factor for SARS-CoV-2 (+) in this patient group.

AIT is recommended for patients who use optimal pharmacological treatment for allergic respiratory diseases, minimize allergen exposure, but do not get enough benefit from these treatments or have side effects related to these treatments. Thus, AIT can be considered as the next step of pharmacological treatment in patients with AR. Therefore, it is expected that the duration of illness in patients with AR who underwent AIT would be longer than in patients without AIT. In patients with a long disease duration, higher serum IgE levels may be achieved due to increased allergen exposure.

The prevalence of SARS-CoV-2, SARS-CoV-2-associated pneumonia, and hospitalizations between patients with and without AIT were found to be normal between groups in our study. In a retrospective study conducted in Wuhan, no difference was found between patients with AR and non-AR patients in terms of severe cases, need for mechanical ventilators, and complications (12). Ren et al. reported that AR has a protective effect for COVID-19 infection in all age groups and that drugs used in the treatment of AR (antihistamines and intranasal steroids) do not affect COVID-19 severity and mortality (13). A study conducted in Turkey showed that there was no significant difference between patients with and without allergic rhinitis in terms of SARS-CoV-2-related hospitalizations and COVID-19 severity (14). It was reported by Vezir et al that COVID-19 is more asymptomatic/mild in pediatric patients with aeroallergen sensitivity (15). A number of hypotheses have been proposed in these studies to explain the relatively positive effect of AR on the course of COVID-19. The first of these is on nasal steroids. Intranasal steroids are the most commonly used drugs in allergic rhinitis patients, In an ARIA-EAACI statement, it has been suggested that patients infected with COVID-19 can use intranasal steroids at recommended doses, since there is no evidence that the immune system is suppressed by these agents, allergic rhinitis errors should not discontinue the use of intranasal steroids. (16). Moreover, some nasal steroids such as mometasone have been shown to inhibit SARS-CoV-2 replication (17). It has been suggested by Straus et al. that the use of nasal steroids reduces COVID-19-related hospitalizations, intensive care admissions, and mortality (18). Another hypothesis is about eosinophils. With experimental studies, eosinophils have been shown to have a potential role in viral clearance and antiviral host defense (19). Clinical prevalence in patients with AR correlates with blood and nasal eosinophil counts (20). Therefore, it can be speculated that increased eosinophil counts in the respiratory tract may be protective against COVID-19 (21). Another hypothesis proposes that ACE2 expression is decreased due to Type 2 inflammation in airway cells of patients with AR, and that allergen-specific T cells show a rapid and effective memory response to heterogeneous SARS-CoV-2 epitopes (13). It has been shown by Kimura et al that IL-13 exposure reduces ACE2 expression in patients with asthma and AR (22). Contrary to these data, Yang et al. suggested that allergic rhinitis has an increased risk in terms of SARS-CoV-2 positivity and more severe disease, and that hospitalizations are longer in patients with allergic rhinitis (23).

As another result of our study, it was found that female gender is protective in terms of SARS-CoV-2 (+) in patients with AR. Many reasons have been suggested in the pathophysiology of the COVID-19 difference between genders (24-26). Many studies have shown since the beginning of the pandemic that male gender is a risk factor for COVID-19-related morbidity and mortality. The reason for these differences between genders may be immunological, hormonal or genetic differences or a combination of these. The effects of sex hormones on pattern recognition receptor and type I IFN responses are different. Sex hormones may affect immune cells in different ways. Estrogen is immunosuppressive at high doses and activates the immune system at low doses. On the other hand, testosterone suppresses natural immunity at all doses. Also, estrogen has been shown to inhibit ACE2, a functional receptor of SARS-COV-2, but androgens upregulate ACE2 activity (27).

As a result of the study, we found that AIT or AIT duration in patients with AR is (neither a risk factor nor protective) for SARS-CoV-2 (+).There is no study in the literature on the course of COVID-19 in patients with allergic rhinitis who underwent AIT. There is increasing evidence that AIT induces IgG4-positive regulatory B cells (Bregs) and regulatory B cells suppress antigen-specific T cell proliferation by producing IL-10. Also, AIT induces Treg cell formation (10, 28). Treg cells are an indispensable subset of T cells to weaken the excessive immune response to pathogens, develop immune tolerance against environmental proteins, cancer cells and transplanted organs, and prevent and control the occurrence of autoimmune and allergic diseases (29). Treg cells can inhibit ongoing inflammation in various steps by secreting suppressive mediators such as IL-10, TGF-β, and IL-35, by suppressing and/or cytolyzing dendritic cells through membrane molecules such as CTLA-4, PD-1, and enzymes such as granzymes A and B (9, 30, 31). T reg cells use these mechanisms to suppress all effector cell types (directly or indirectly), eosinophils, B cells, DCs, T cells as well as inflamed resident tissue cell (29). Therefore, despite the fact that it is thought that AIT can contribute positively to the course of COVID-19 by preventing the exaggerated cytokine response via Treg, we could not reach such a result in our study and we found the COVID-19 course of patients with AR who underwent AIT and patients with AR who did not receive AIT to be similar. We believe that more comprehensive studies should be conducted on this subject.

Our study has some limitations. The first of these is the cross-sectional design. Secondly, the age range of the study population is much younger compared to older patients who are vulnerable to COVID-19, and they have fewer accompanying comorbid non-allergic comorbidities. Another thing that may have negatively affected the prevalence of SARS-CoV-2 is that rapid diagnostic tests such as PCR were not widely used at the beginning of the Pandemic.

In conclusion, we would like to highlight that the course of COVID-19 is similar in patients with AR who underwent AIT and patients with AR who did not undergo AIT, and AIT does not seem to increase the risk for COVID-19 infection. So, It could be safely use in patients with AR, compatible with the data in the literature.

## Data Availability

All data produced in the present study are available upon reasonable request to the authors

## Acknowledgement

None

